# Neighborhood Deprivation Is Associated with Accelerated Epigenetic Aging Via Greater Individual Adversity

**DOI:** 10.64898/2026.04.24.26351669

**Authors:** Alija Koirala, Justin R. Shields, Anjali S. Vijan, Stephanie Wemm, Ke Xu, Benson S. Ku, Rajita Sinha, Zachary M. Harvanek

**Affiliations:** Department of Psychiatry, Yale University School of Medicine, New Haven, CT; Department of Psychiatry and Behavioral Sciences, Emory University School of Medicine

## Abstract

**Importance:** Adverse neighborhood conditions can lead to poorer health outcomes, potentially through accelerated biological aging. However, whether these relationships are explained by individual- or neighborhood-level factors remains unclear.

**Objective:** To examine the association between neighborhood deprivation, measured by the Area Deprivation Index (ADI), and epigenetic age acceleration and assess whether individual- and neighborhood-level characteristics mediate or modify these associations.

**Design:** Cross-sectional study using data from a Yale Stress Center study between 2008 and 2012. Data analysis was conducted from July 2025 to January 2026.

**Setting:** Community-based sample from the greater New Haven, CT area.

**Participants:** A total of 370 healthy adults aged 18 to 50 years without major psychiatric, medical, or cognitive disorders who provided blood samples for DNA methylation analysis.

**Main Outcomes and Measures:** Epigenetic age acceleration measured from DNA methylation using four second-generation epigenetic clocks, with associations assessed among aging, neighborhood deprivation, and individual- and neighborhood-level factors.

**Results:** Data were analyzed from 370 participants (212 women [57.3%], 158 men [42.7%]; mean [SEM] age, 29.3 [0.46] years). Greater neighborhood deprivation was associated with greater lifetime adversity (β=0.112, p<.001) and lower educational attainment (β=-0.019, p=.012), and accelerated epigenetic aging as measured by GrimAge (β=0.037, p<.001), PCGrimAge (β=0.019, p<.001), and PCPhenoAge (β=0.041, p<.001), but not PhenoAge (p=.23). In multivariable models accounting for individual factors, neighborhood deprivation remained associated with these three clocks. Lifetime adversity partially mediated the association between ADI and accelerated GrimAge (20.3% of total effect) and PCGrimAge (23.3%). Race moderated the direct association between ADI and epigenetic aging, with stronger associations between neighborhood deprivation and accelerated GrimAge (β=0.061, p=.004) and PCPhenoAge (β=0.057, p=.02) observed among Black participants compared to White.

**Conclusions:** Greater neighborhood deprivation was associated with accelerated epigenetic aging across multiple second-generation clocks, with lifetime adversity partially mediating these associations. Stronger effects were observed among Black participants. These findings suggest that neighborhood environments and cumulative stress may contribute to biological aging and racial disparities in aging trajectories.

**Key Points:** *Question:* Is neighborhood deprivation associated with epigenetic age acceleration, and if so, how do neighborhood- and individual-level factors impact this relationship?

*Findings:* In this cross-sectional study of 370 adults, greater neighborhood deprivation was associated with accelerated epigenetic aging across multiple second-generation clocks. Lifetime adversity partially mediated these associations, and the relationship between neighborhood deprivation and accelerated aging was stronger among Black participants than White participants.

*Meaning:* These findings suggest that neighborhood conditions and lifetime stress contribute to accelerated biological aging and suggest that epigenetic aging may represent one biological pathway through which neighborhood-level racial inequalities contribute to health disparities.

## Introduction

Neighborhood environments are powerful determinants of health and aging trajectories. Residents of high deprivation neighborhoods are disproportionately exposed to environmental pollution, chronic stress, have limited access to nutritious food, and fewer education and employment opportunities^1^. These structural disadvantages can have direct and cumulative health effects, including poorer psychological health, increased risk of diseases like type 2 diabetes, and heightened vulnerability to other age-related conditions^2–4^. Collectively, these burdens translate into higher mortality and societal costs. In extreme cases, residents in disadvantaged neighborhoods can live up to 15 years shorter than residents of more affluent neighborhoods within the same city^5^. While the biological mechanisms linking neighborhoods to health and mortality remain unclear, emerging evidence suggests that neighborhood-level deprivation may accelerate biological aging though changes in cellular and molecular markers of aging^6,7^.

Epigenetic mechanisms, such as DNA methylation, modify gene expression without altering the DNA sequence and are highly sensitive to environmental exposures^8^. DNA methylation-based “epigenetic clocks” estimate biological age using methylation patterns across CpG sites to predict aging and aging-related phenotypes^9,10^. First-generation clocks were trained to predict chronological age, whereas second-generation clocks such as GrimAge and PhenoAge were designed to predict age-related morbidity and mortality and demonstrate superior associations with these outcomes^11^. Higher levels of individual-level adversity and trauma have been associated with accelerated aging measured by these second-generation clocks^12,13^. Notably, prior work has also demonstrated associations between cumulative neighborhood disadvantage and second-generation epigenetic clocks among urban Black adults, underscoring the potential role of structurally patterned environmental exposures^14^.

One widely used measure of neighborhood deprivation is the Area Deprivation Index (ADI), a census-based socioeconomic index that integrates indicators of income, education, employment and housing quality to rank neighborhoods by disadvantage^15^. Higher ADI scores have been associated with worse health outcomes, including cardiovascular disease^16^, dementia^17^, premature mortality^18^, and greater healthcare utilization^19^. Recent evidence also links higher ADI scores to accelerated epigenetic aging in specific populations, such as women who had a sister with breast cancer^20^ or trauma-exposed individuals^21^. Despite the ADI’s strong predictive validity of adverse health outcomes, neighborhood-level measures remain underutilized in the social epigenetics literature, which often focuses on a specific demographic group and assesses deprivation primarily on the individual or household level^22^. This focus may miss potential crosstalk between individual-level and neighborhood-level pathways affecting biological aging. Moreover, most prior studies demonstrating associations between ADI and epigenetic aging have not jointly modeled individual-level adversity alongside neighborhood deprivation, nor examined the potential mediation and moderation of these relationships^14,20^.

To address this gap, we examined how individual-level and neighborhood-level adversity factors relate to biological aging as measured by second-generation epigenetic clocks. Specifically, we (1) tested whether ADI and specific neighborhood and individual-level characteristics are associated with epigenetic aging; (2) assessed whether these associations persist after mutual adjustment for individual-and neighborhood-level factors; (3) conducted exploratory mediation analyses to evaluate whether individual-level lifetime adversity and education mediate the ADI-epigenetic aging relationship; (4) and tested whether race moderates these pathways using moderated mediation models.

## Methods

### Cohort

This study was conducted utilizing data from a Yale Stress Center cohort recruited between 2008 to 2012 examining the role of stress, emotion regulation, and self-control^23^ (see Supplementary Methods). The current study included 370 healthy community-recruited adults aged 18-50 years in the greater New Haven, CT area for whom both epigenetics and address were available. All participants provided verbal and written informed consent, and the study protocol was approved by the Yale Institutional Review Board (IRB).

### Individual-Level Variables

Sociodemographic and health-related variables were collected during the participant’s baseline interview, including race, years of education, lifetime adversity, and monthly income from any source. Lifetime adversity was assessed using the Cumulative Adversity Inventory (CAI), a 140-item interview-based assessment of stressful life events, such as work, financial, traumatic, relationship, family and neighborhood- related stressors across the lifetime, as well as participants’ perceived sense of being overwhelmed by specific events^24^. Higher scores indicate a higher overall level of lifetime adversity.

### Neighborhood-Level Variables

Neighborhood-level variables were derived from geocoded residential addresses using U.S. Census and geospatial datasets. Data extraction was performed in R via a custom geocoding script originally written by Dr. Ku and adapted by Dr. Harvanek. Data were derived from the 2010 American Community Survey (ACS) 5-year estimates^25^, USDA Neighborhood Food Access Atlas^26^, the Area Deprivation Index (ADI) from the Neighborhood Atlas^15^, NOD estimates from Annual Land Use Regression Model Estimates^27^, and the Historic Redlining Indicator^28^. ADI was modeled continuously as a national percentile rank (1-100), with higher scores indicating greater deprivation. Residential addresses at study enrollment were linked to the 2015 ADI (the closest publicly available data to the recruitment timeframe) using Census block group identifiers.

Neighborhood-level indicators were selected to capture socioeconomic conditions, racial composition, and environmental exposures relevant to aging disparities. Indicators included the Area Deprivation Index (ADI), neighborhood education (percentage of residents with ≥4 years of college vs. <4 years), annual neighborhood income (median household income ≥$100,000 vs. <$100,000), percentage of White residents, historical redlining status, pollution exposure (annual mean NOD concentration), and food access (proportion of housing units without a vehicle and low supermarket access).

### Clock Calculation

Genomic DNA for epigenetic analysis was isolated from whole blood samples^14^. Briefly, DNA methylation was quantified using Illumina Infinium Human Methylation450 Beadchip, with quality control as previously published^12^. Epigenetic age was estimated using the MethylCIPHER package in R^29^. Mortality-trained clocks GrimAge and PhenoAge^30,31^ were calculated by both traditional and principal-component based methods^11^. Epigenetic age acceleration was defined as the residuals obtained from regressing epigenetic age onto chronological age. Cell-type proportions were estimated using the Houseman method^32^.

### Statistical Analyses

Analyses were performed using R version 4.0.2^33^. Associations among epigenetic aging, psychological factors, and neighborhood-level variables were assessed with linear mixed-effects models using the lmerTest package^34^, with random intercepts for census tracts. When mixed-effects models failed to converge (in analyses involving associations between neighborhood variables and ADI), we used standard linear regression models.

Mediation and moderated mediation models were estimated using structural equation modeling in lavaan, described in detail in the supplementary methods. Briefly, neighborhood disadvantage (ADI rank) was specified as the predictor, lifetime adversity and years of education as parallel mediators, and epigenetic aging indices as outcomes, adjusting for age, gender, and immune cell composition. Race (Black=1, White=0) was tested as a moderator of a-, b-, and/or c′-paths via interaction terms; only significant moderation pathways were retained for parsimony.

For models examining epigenetic age acceleration, multivariable regressions were adjusted for sex, age, and cell-type proportions (dropping granulocytes to avoid overfitting). Unstandardized β-values are presented in text and tables unless otherwise specified. Figures display standardized coefficients to facilitate comparison across predictors and clocks. All statistical tests were two-tailed with _α_=0.05. Nominal p-values are reported for consistency. For linear and mixed-effects models examining epigenetic clocks (e.g., figures 1 and 2), we correct for multiple comparisons using the Bonferroni method (nominal significance threshold p<0.0125, corresponding to _α_=0.05). As mediation analyses were performed as specific exploratory follow-up analyses, no correction was used for these models.

**Figure 1:**
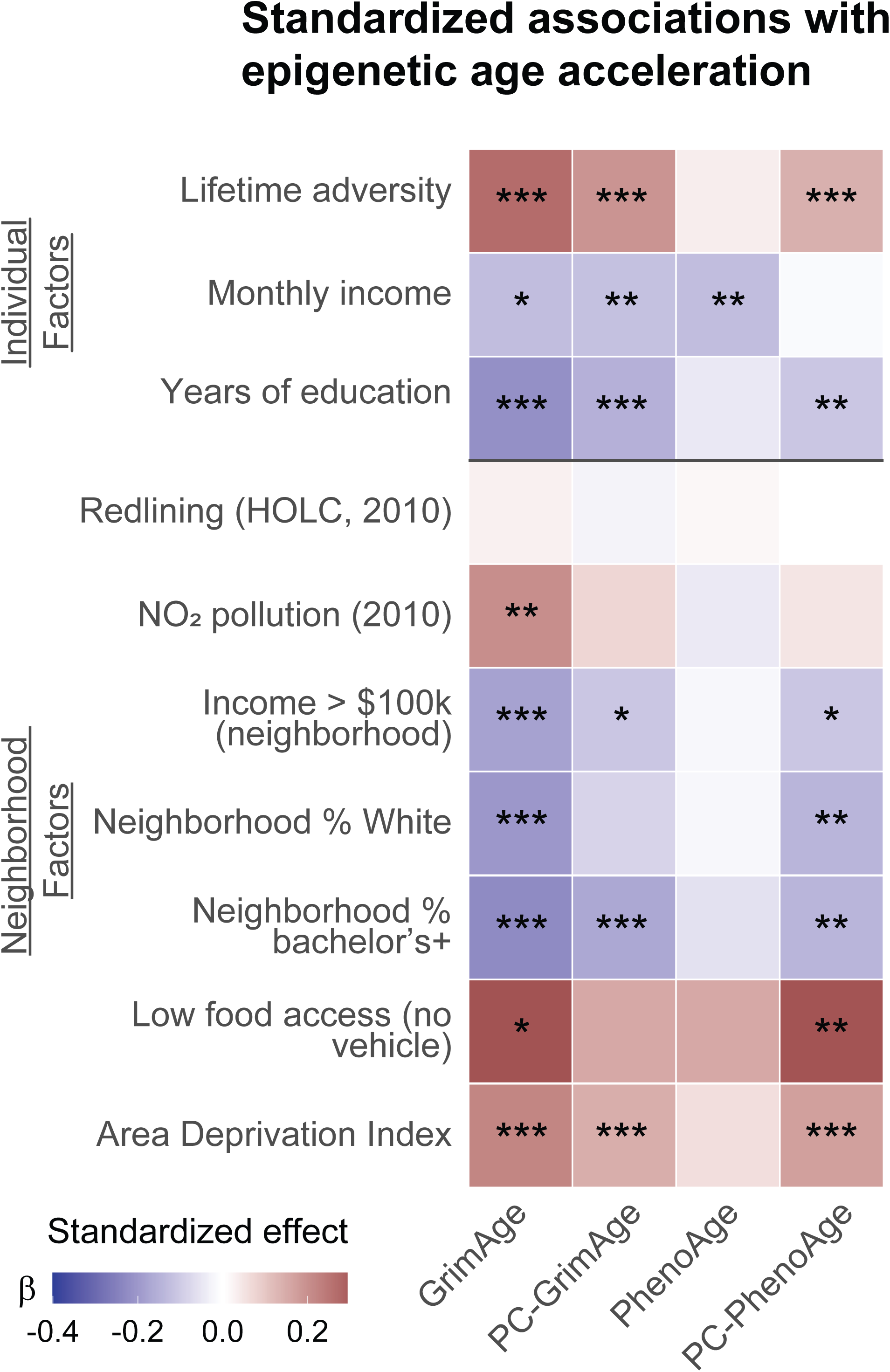
Associations between individual and neighborhood factors and epigenetic aging. Heatmap showing association between individual or neighborhood factors and epigenetic aging in linear mixed-effects models with random intercept for census tract. Adjusted for Age, Gender, CD4T, CD8T, Mono, NK, Bcell. ß: SD change in outcome per 1 SD in predictor (numeric) or group difference (factors). Tiles show standardized effects (ß). Satterthwaite p-values: *p < 0.05 (nominal significance); **p < 0.0125 (Bonferroni-corrected significance across 4 tests); *** p<0.001. Outcome & continuous covariates standardized; Gender and food access are treated as factors.

## Results

### Demographics and clinical characteristics

Our study population (n=370) were healthy community adults without chronic medical or psychiatric diseases (Table 1). Most participants were female (57.3%), non-smokers (79.5%), and identified as White (72.3%).

**Table 1:**
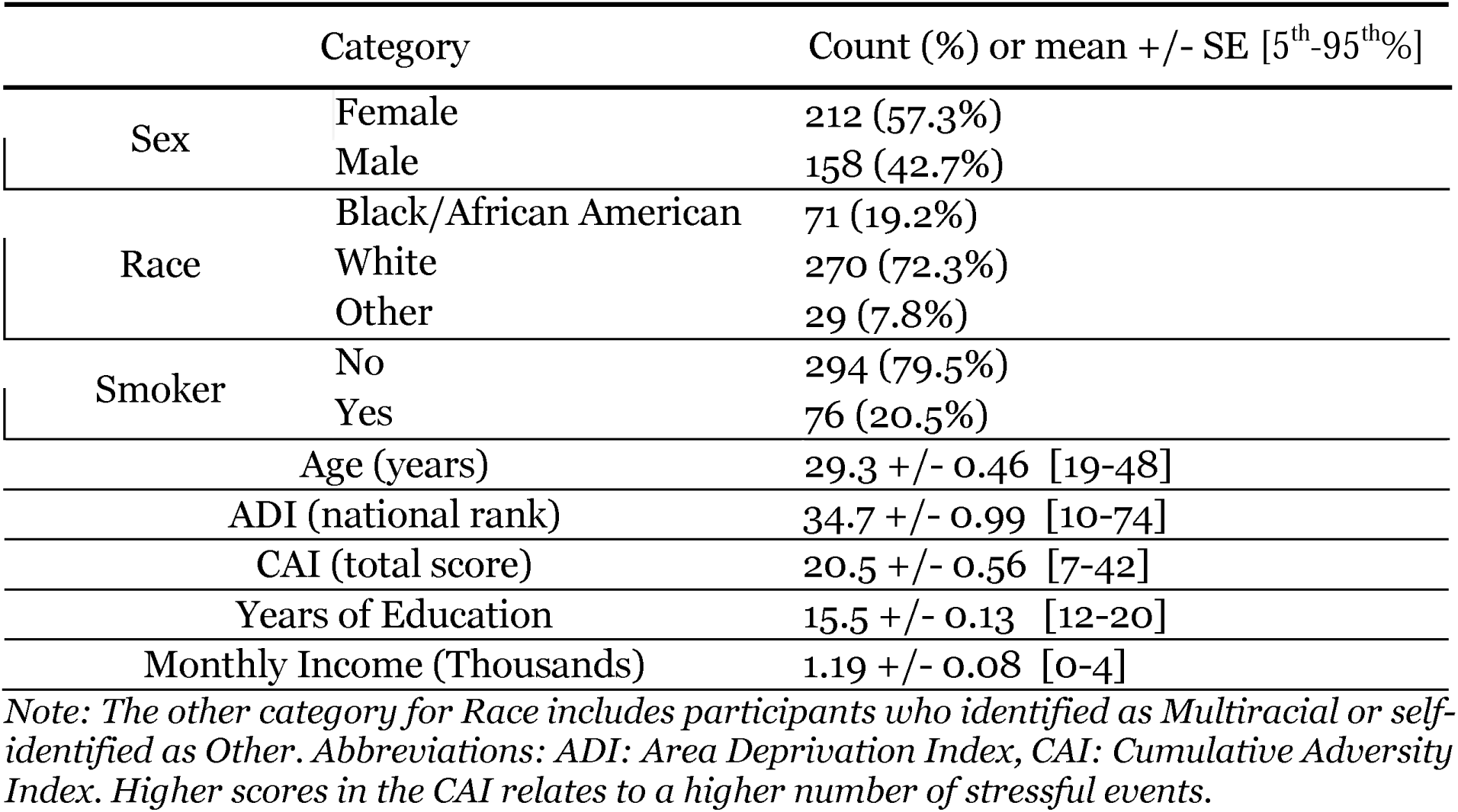
Characteristics of study participants.

|  | Category | Count (%) or mean +/- SE [5<sup>th</sup>-95<sup>th</sup>%] |
| --- | --- | --- |
| Sex | Female | 212 (57.3%) |
|  | Male | 158 (42.7%) |
| Race | Black/African American | 71 (19.2%) |
|  | White | 270 (72.3%) |
|  | Other | 29 (7.8%) |
| Smoker | No | 294 (79.5%) |
|  | Yes | 76 (20.5%) |
| Age (years) |  | 29.3 +/- 0.46 [19-48] |
| ADI (national rank) |  | 34.7 +/- 0.99 [10-74] |
| CAI (total score) |  | 20.5 +/- 0.56 [7-42] |
| Years of Education |  | 15.5 +/- 0.13 [12-20] |
| Monthly Income (Thousands) |  | 1.19 +/- 0.08 [0-4] |

*Note: The other category for Race includes participants who identified as Multiracial or self-identified as Other. Abbreviations: ADI: Area Deprivation Index, CAI: Cumulative Adversity Index. Higher scores in the CAI relates to a higher number of stressful events.*

### Univariate associations between individual and neighborhood factors and epigenetic age acceleration

#### Associations Between ADI, Individual Factors, and Neighborhood Factors

We first assessed if ADI was associated with individual-level measures of adversity and socioeconomic disadvantage. Higher ADI was associated with greater lifetime adversity and lower educational attainment (Table 2), but not individual income (p=.23).

**Table 2:** Associations between ADI and Individual/Neighborhood-level Factors and Epigenetic Clocks.

| Outcomes | $\beta$ | *p* -value | Standard Error |
| --- | --- | --- | --- |
| **Individual-Level Factors** |  |  |  |
| Lifetime Adversity | 0.112 | <.001 | 0.033 |
| Educational Attainment | -0.019 | .012 | 0.0076 |
| Individual Income | -4.002 | .360 | 4.352 |
| **Neighborhood-Level Factors** |  |  |  |
| *NO*<sub>2</sub> Exposure | 0.032 | <.001 | 0.0073 |
| Neighborhood Income | -0.64 | <.001 | 0.029 |
| Historical Redlining | 0.0095 | <.001 | 0.0024 |
| Neighborhood Education | - 0.64 | <.001 | 0.046 |
| % White Residents | -0.97 | <.001 | 0.05 |
| Food Access | 0.013 | <.001 | 0.00095 |

| **Epigenetic Aging Clocks** |  |  |  |
| --- | --- | --- | --- |
| GrimAge Acceleration | 0.037 | <.001 | 0.0084 |
| PCGrimAge Acceleration | 0.019 | <.001 | 0.0058 |
| PhenoAge Acceleration | 0.0173 | .23 | 0.014 |
| PCPhenoAge Acceleration | 0.041 | <.001 | 0.01 |

*Note: Table shows univariate associations between ADI and individual factors, neighborhood factors, and epigenetic clocks. Individual-level and epigenetic clocks use linear mixed-effects models accounting for census tracts. Neighborhood factors used standard linear regression due to lack of convergence.*

We next examined relationships between ADI and neighborhood factors. As expected, higher ADI was associated with more adverse neighborhood conditions, including greater NO□ exposure, lower neighborhood income, greater historical redlining, lower neighborhood educational attainment, a lower percentage of white residents in a neighborhood, and reduced food access. β coefficients, standard errors, and p values for all models are presented in Table 2.

#### Associations Between ADI and Epigenetic Aging

Higher ADI was linked to accelerated aging as measured by GrimAge (β=0.037), PCGrimAge (β=0.019), and PCPhenoAge (β=0.041) (see figure 1, all ps<.001), but not with PhenoAge (p=.135) in models accounting for age, sex, and cell proportions. Results were consistent in sensitivity analyses either excluding cell proportions (eTable 1) or including individual smoking status as a covariate (GrimAge/PCGrimAge/PCPhenoAge: ps<0.001, PhenoAge: p=0.21).

#### Associations between Individual Factors and Epigenetic Aging

We next examined associations between individual factors and epigenetic aging (summary in Figure 1, detailed estimates provided in eTable 2). After adjusting for age, sex, and cell proportions, lower individual education and greater lifetime adversity were associated with accelerated GrimAge (education: β=−0.27, adversity: β=0.080), PCGrimAge (β=−0.15, β=0.045), and PCPhenoAge (β=−0.19, β=0.060), but not PhenoAge (education: p=.32; adversity: p=.47).

Lower individual income was associated with accelerated PCGrimAge and PhenoAge (β=−0.00020, β=−0.00047), and nominally with GrimAge (β=−0.00027), but not PCPhenoAge (p=.72).

#### Neighborhood Factors and Epigenetic Aging

Neighborhood-level associations with aging are summarized in Figure 1 (details in eTable 2). Lower neighborhood education and income showed generally similar patterns. Neighborhood education was associated with accelerated GrimAge (β=−0.035), PCGrimAge (β=−0.021), and PCPhenoAge (β=−0.031), while neighborhood income was associated with GrimAge (β=−0.035) and, nominally, with PCGrimAge (β=−0.017) and PCPhenoAge (β=−0.030).

Lower percentage of white residents was associated with accelerated GrimAge (β=−0.025) and PCPhenoAge (β=−0.025). Lower food access was associated with accelerated PCPhenoAge (β=1.4) and, nominally, GrimAge (β=1.0). Greater NOD exposure was associated only with accelerated GrimAge (β=−0.80) Historical redlining was not associated with any epigenetic aging measures (all ps > .05).

### Multivariable associations between individual and neighborhood factors and epigenetic age acceleration

#### Associations between Epigenetic Clocks and ADI, accounting for individual factors

Next, we examined whether associations between ADI and epigenetic aging were independent of individual-level variables (Summary in figure 2A, details in eTable 3).

**Figure 2:**
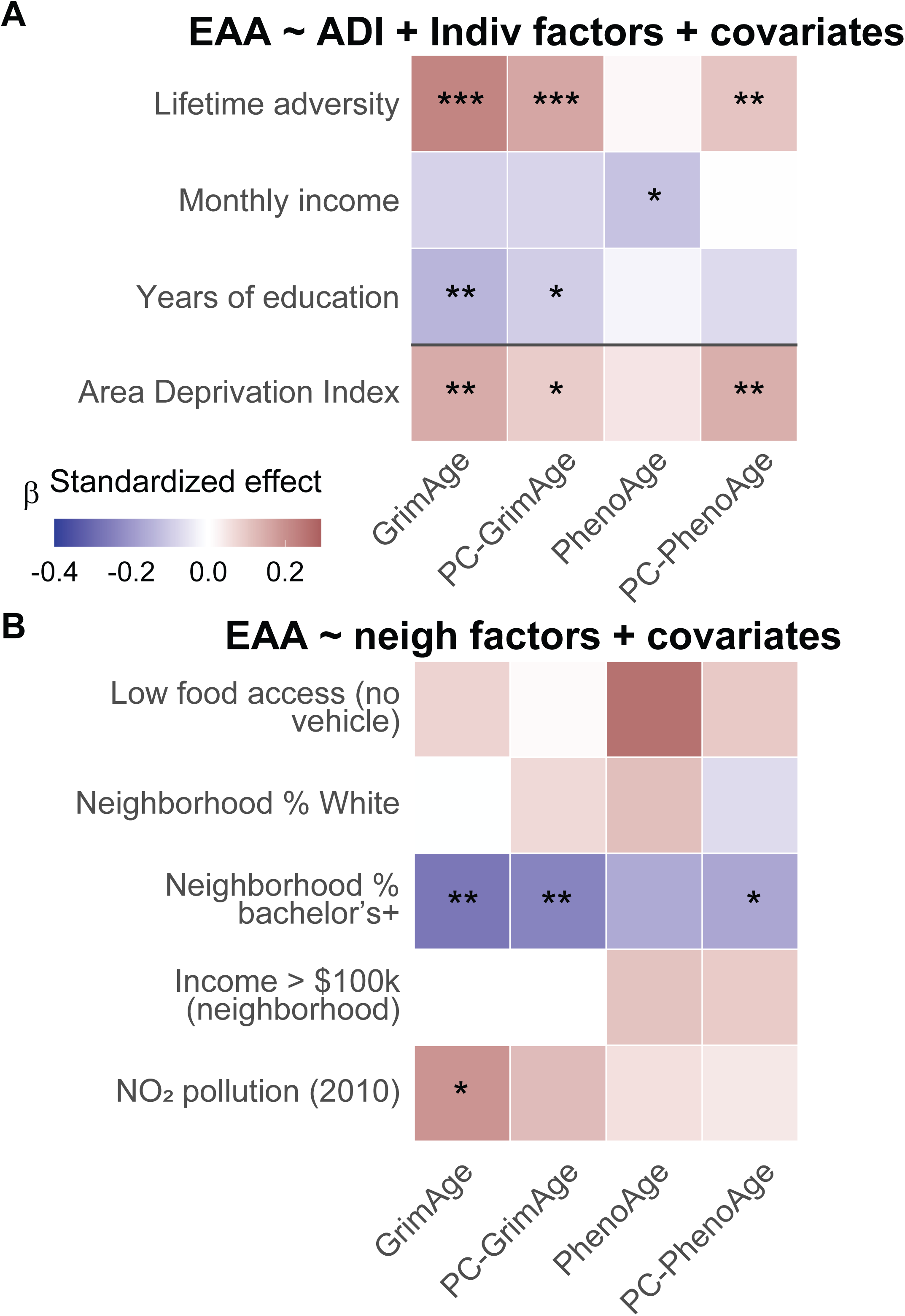
Multivariable models demonstrate effects of ADI on epigenetic aging are not fully accounted for by individual factors. Heatmap showing association between ADI and epigenetic aging accounting for either individual (A) or neighborhood (B) factors in multivariable linear mixed-effects models with random intercept for census tract. Adjusted for Age, Gender, CD4T, CD8T, Mono, NK, Bcell. ß: SD change in outcome per 1 SD in predictor (numeric) or group difference (factors). Tiles show standardized effects (ß). Satterthwaite p-values:. *p < 0.05 (nominal significance); **p < 0.0125 (Bonferroni-corrected significance across 4 tests); *** p<0.001. Outcome & continuous covariates standardized; Gender and food access are treated as factors.

In multivariable mixed-effects models accounting for age, sex, cell proportions, and individual factors, higher neighborhood deprivation (ADI) remained associated with accelerated GrimAge (β=0.025), PCPhenoAge (β=0.034), and nominally associated with PCGrimAge (β=0.012). Greater lifetime adversity showed significant associations in the same clocks as in earlier models: GrimAge (β=0.066), PCGrimAge (β=0.038), and PCPhenoAge (β=0.044). Individual education predicted decelerated GrimAge (β=−0.17) and, nominally, PCGrimAge (β=−0.094) but not PCPhenoAge (p=.072). Individual income only showed a nominal association with PhenoAge (β=-0.00043) but no longer showed significant associations with GrimAge (p=.078) or PCGrimAge (p=.07).

In sensitivity analyses adjusting for individual smoking in addition to individual education, income, and lifetime adversity, ADI remained associated with GrimAge (β=0.027, p<0.001), PCGrimAge (β=0.014, p=0.007), and PCPhenoAge (β=0.035, p=0.001).

#### Associations between Epigenetic Clocks and Neighborhood Factors

We next asked whether specific neighborhood-level factors have independent effects on epigenetic aging. In multivariable mixed-effects models adjusting for age, sex, and cell proportions, and all neighborhood variables (except ADI) that showed significant effects on any clock in prior analyses (Figure 2B, full estimates in eTable 4), higher neighborhood educational attainment was associated with decelerated GrimAge (β=−0.041, p=0.0083), PCGrimAge (β=−0.030, p=0.0042), and, nominally, PCPhenoAge (β=−0.038, p=0.038). Greater NOD exposure was nominally associated with accelerated GrimAge (β=0.24, p=0.036). Neighborhood income, racial composition, and food access were not independently associated with epigenetic aging in any clocks.

### Mediating roles of individual factors in the ADI-Epigenetic Age Acceleration Relationship

Given attenuation of ADI associations with epigenetic aging in multivariable models, we tested whether some individual-level factors mediated associations between ADI and epigenetic aging (full details for mediation analyses in eTable 5 in Supplement 1). In mediation models adjusting for age, sex, and cell type proportions, neighborhood deprivation (ADI) was positively associated with GrimAge acceleration, through both direct and indirect pathways. Lifetime adversity partially mediated this association, accounting for approximately 20.3% of the total effect of ADI on GrimAge acceleration (indirect effect: p=.030, Figure 3A). The indirect effect through education was not significant (p=.073). ADI continues to have a significant direct effect on GrimAge Acceleration (p=.002).

**Figure 3:**
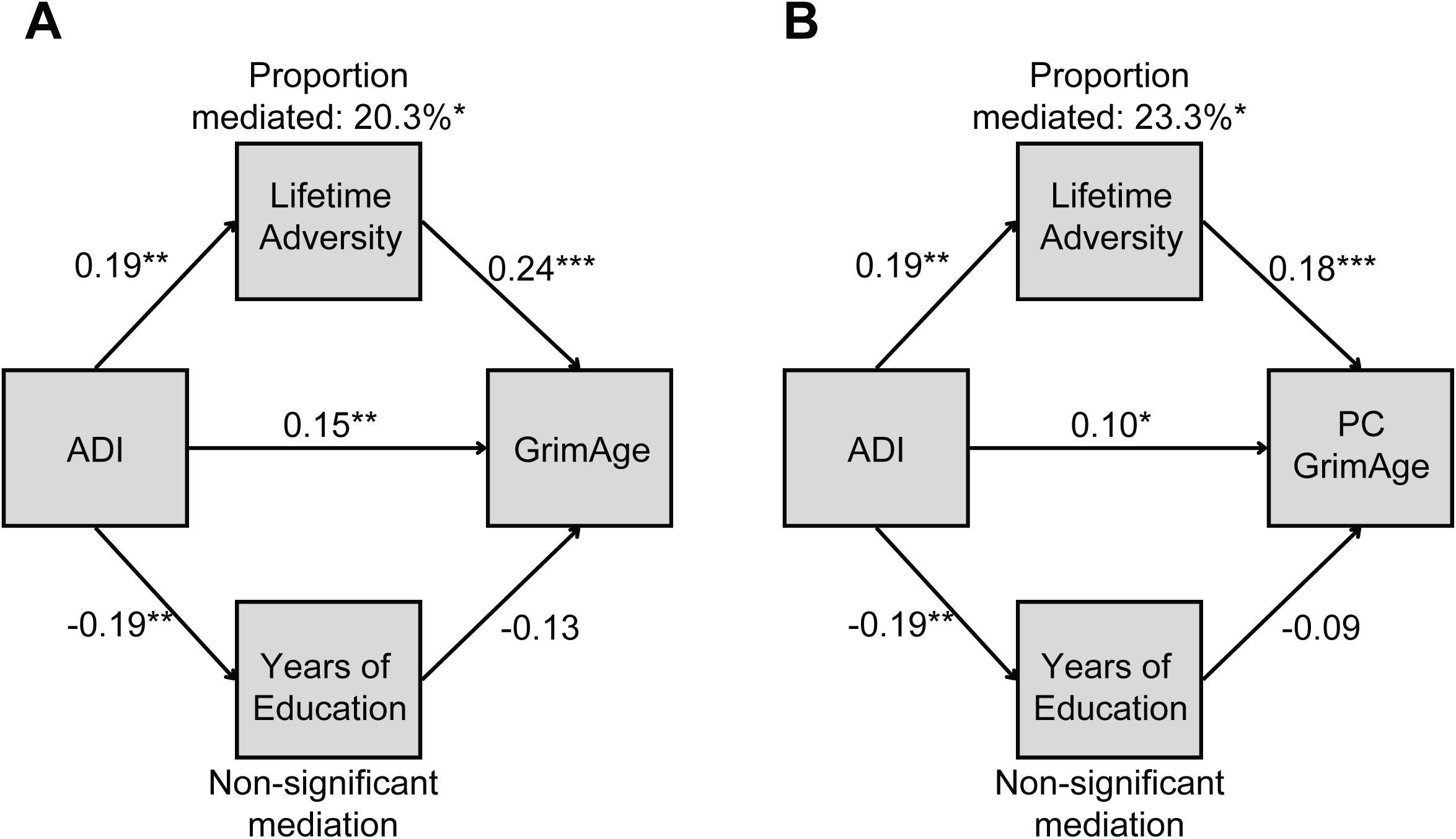
Mediation of the Association between Neighborhood Deprivation and Epigenetic Aging by Lifetime Adversity. Panels A-B A: Lifetime adversity significantly mediated the association between ADI and GrimAge acceleration, accounting for 20.3% of the total. B: Lifetime adversity significantly mediated the association between ADI and PCGrimAge acceleration, accounting for 23.3% of the total. For both panels, mediation models examine lifetime adversity and individual education as potential mediators of the relationship between neighborhood deprivation (as measured by ADI) and epigenetic aging outcomes. Figures show models with GrimAge and PCGrimAge. Numbers represent standardized effect sizes. Asterisk indicates significance (* p < .05, ** p < 0.01, *** p < 0.001) as determined via bootstrapping (n = 5000 iterations). Proportion mediated indicates the % of the total effect mediated through lifetime adversity. Years of education were not a significant mediator for either model.

Similar effects were observed for PCGrimAge (indirect effect via lifetime adversity: p=.034, Figure 3B). Lifetime adversity accounted for 23.3% of the total effect. Education was not a significant mediator.

No significant mediation was observed for PCPhenoAge (lifetime adversity: p=0.091, individual education: p=0.068), although ADI remained directly associated with PCPhenoAge (p=0.001).

No significant indirect or direct effect of ADI on PhenoAge was observed.

### Moderating roles of individual and neighborhood factors in the ADI-Epigenetic Age Acceleration Relationship

As prior findings have demonstrated race-related differences between Black and White individuals in both epigenetic aging and neighborhood deprivation^35^, we next examined whether race moderated associations between ADI and epigenetic aging, including individual race as a moderator of all pathways, as well as a direct effect of race on epigenetic age (full estimates reported in eTable 6).

Race only showed significant moderating effects on the direct association between ADI and GrimAge and PCPhenoAge acceleration, with stronger associations between ADI and accelerated aging observed among Black participants. Because moderation was limited to the direct ADI-aging association, final models included race as a moderator of the direct pathway only and focus on GrimAge and PCPhenoAge.For GrimAge, both the total and direct effects of ADI differed significantly by race (p=.004, Figure 4A, Full estimates in eTable 7), with neighborhood deprivation leading to more age acceleration in Black individuals.

**Figure 4:**
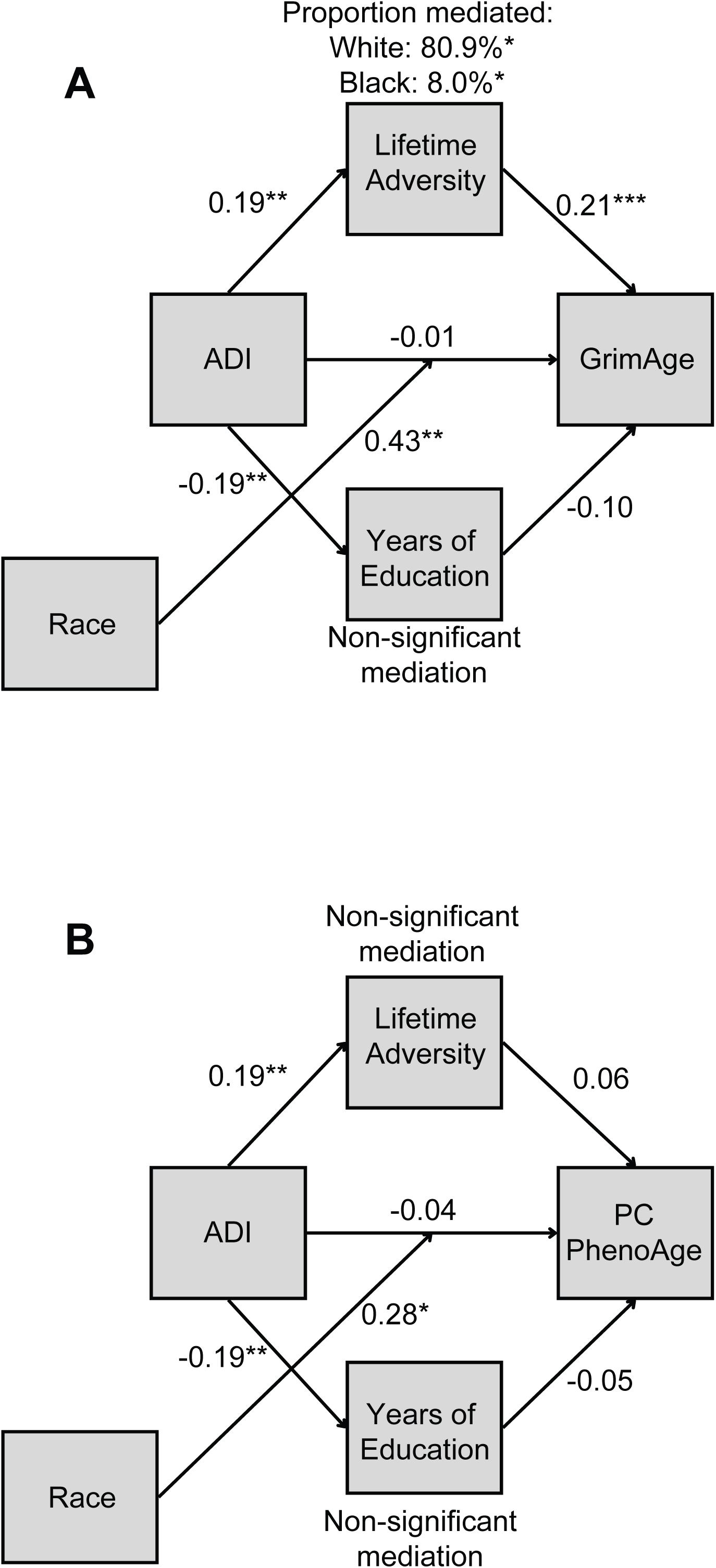
Moderated Mediation of the Association between Neighborhood Deprivation and Epigenetic Aging by Race. A: The association between ADI and GrimAge acceleration is different by race (p = 0.004), with lifetime adversity mediating 8.5% of the total effect (80.9% in White participants, 8.0% in Black participants). B: Race moderated the association between ADI and PCPhenoAge acceleration, but lifetime adversity was not a significant mediator. For both panels, moderated mediation models examine whether race modifies the direct association between neighborhood deprivation (as measured by ADI) and epigenetic aging, with lifetime adversity and individual education remaining as potential mediators of the relationship. Figures 4A and 4B show the model with GrimAge and PCPhenoAge. Numbers represent standardized effect sizes. Asterisk indicates significance (* p < .05, ** p < 0.01, *** p < 0.001) as determined via bootstrapping (n = 5000 iterations). Proportion mediated indicates the % of the total effect mediated through lifetime adversity. Years of education were not a significant mediator in either model, and lifetime adversity lost significance in the PCPhenoAge model. Due to sample size limitations, analyses for race only include White (coded as 0) and Black (coded as 1) individuals.

Lifetime adversity continued to partially mediate this association (indirect effect: p=.033), accounting for 80.9% of the effect in White participants, and 8% in Black participants. This difference is driven by a significantly smaller direct effect of ADI on GrimAge in White compared to Black participants. Education was not a significant mediator.

For PCPhenoAge, we observe a similar moderating effect, with higher neighborhood deprivation leading to significantly higher age acceleration in Black compared to White individuals (p=.02, Figure 4B). Neither lifetime adversity (p=0.214) nor individual education (p=0.227) significantly mediated this association.

## Discussion

This study examined how both individual-level and neighborhood-level factors together affect epigenetic aging in second-generation clocks. We identify associations between multiple neighborhood deprivation factors and increased epigenetic age across multiple second-generation clocks, which were most consistently observed with the ADI and neighborhood education levels. Neighborhood effects remain significant even after accounting for individual-level adversity, income, and education. Lifetime adversity partially mediates the relationship between ADI, GrimAge, and PCGrimAge. Notably, race moderates the direct effects of ADI on GrimAge and PCPhenoAge, suggesting neighborhood deprivation may interact with societal factors such as structural racism. Overall, these findings suggest a complex interplay between neighborhood-level and individual-level factors, with both contributing uniquely and in tandem to increased epigenetic age.

### Both Individual and Neighborhood Variables Play a Role

Neighborhood deprivation (ADI) was associated with greater lifetime adversity, lower individual educational attainment, and all adverse neighborhood characteristics. Higher ADI was also associated with acceleration of multiple clocks, these associations were robust to sensitivity analyses even when accounting for lifetime adversity and educational attainment, supporting an independent contribution of neighborhood deprivation on epigenetic aging.

To examine which neighborhood characteristics may independently contribute to aging, we evaluated specific neighborhood variables. The most consistent results were observed with regard to neighborhood educational attainment, though greater NOD exposure was also nominally associated with accelerated GrimAge. In combination with the finding that individual-level education was associated with GrimAge and (nominally) PCGrimAge, these findings suggest that education may be a key intervention point for both individual and community health.

Prior work has separately linked both individual characteristics (i.e. lower education, socioeconomic status, and cumulative lifetime adversity^12,13,36,37^) and neighborhood characteristics (i.e. pollution and poverty^38,39^) to accelerated epigenetic aging. Some studies have begun to incorporate both individual and neighborhood factors^40^, although neighborhood context is often limited to only neighborhood SES^10^. A recent study in trauma-exposed individuals examined the association between ADI and epigenetic aging and found that individual psychosocial factors (CD-RISC) moderated this relationship^21^. Our findings extend these observations to a community-based sample without recent trauma exposure or diagnosed PTSD and suggest that the influence of neighborhood and individual factors on aging may extend across different population contexts.

### Few Associations with the PhenoAge Epigenetic Clock

Notably, many individual and neighborhood-level measures, including ADI, showed at least nominal associations with three of the four epigenetic clocks, though the traditional version of PhenoAge showed comparatively few significant associations. PhenoAge’s low correlations with individual and neighborhood-level adversity factors suggest that GrimAge and principal component-based measures better capture the social and environmental influences on epigenetic aging. Prior work supports this distinction: Lawerence et al (2020)^20^ found associations between neighborhood deprivation and both PhenoAge and GrimAge, though effect sizes were higher and findings were more robust with GrimAge. A study using the Health and Retirement Study found similar effect sizes of neighborhood factors on PCPhenoAge and PCGrimAge^41^. In another cohort examining individual-level adversity, we previously found associations between childhood adversity and GrimAge, PCPhenoAge, and PCGrimAge, but not PhenoAge^42^. Xu et al (2025) also reported that racial disparities were detectable using GrimAge but not PhenoAge, suggesting greater sensitivity of GrimAge to biological pathways shaped by racial, psychosocial, and structural disadvantages^14^.

### Lifetime Adversity: A Significant Mediator

Mediation analyses showed that lifetime adversity partially explained the association between ADI and accelerated GrimAge and PCGrimAge. Individual education did not independently mediate these relationships. These findings suggest a stress-associated model of accelerated epigenetic aging, in which neighborhood deprivation may contribute to increased life stress exposure which, potentially via altered glucocorticoid signaling or other mechanisms, may alter DNA methylation^13^. Our mediation analyses suggest that individuals in more deprived neighborhoods also experience higher adversity, which in turn results in accelerated aging. Although reverse causation (e.g., higher levels of adversity leading to residence in more deprived areas) is possible, our results are consistent with prior work linking cumulative stress and trauma to accelerated epigenetic aging^13,36,43,44^.

### Structural Racism

In moderated-mediation analyses, we examine individual race as a proxy for unmeasured social and personal factors, which could include structural racism, discrimination, differential policing, barriers to healthcare access, and cumulative exposures to racialized and economic injustice. Race moderated the relationship between neighborhood deprivation and epigenetic aging for both GrimAge and PCPhenoAge, such that the association was stronger in Black individuals as compared to White individuals. The difference was particularly stark in analyses of GrimAge, which suggested that over 80% of the effect of neighborhood deprivation on White participants was through higher levels of adversity, whereas adversity only mediated 8% of the effects of neighborhood deprivation on aging in Black individuals.

One possibility is that these differences could reflect a failure of the CAI to identify types of adversity particularly relevant to Black Americans, such as discrimination and microaggressions. Future work should specifically examine the role of these individual stressors in the relationship between neighborhood and aging. Beyond measurement issues, this pattern also suggests that structural factors correlated with race not fully captured by ADI or other measured variables may amplify the negative effects of neighborhood deprivation. Consistent with race as a proxy for structural racism, existing studies demonstrate racial differences in epigenetic age acceleration linked to living environments. Krieger et al (2024)^45^ reported epigenetic age acceleration associated with birth in a Jim Crow state or residence in highly segregated, low-income Black neighborhoods, and Xu et al (2025)^14^ found faster epigenetic aging among Black adults compared to White adults across multiple clocks.

Structural inequalities such as segregation and divestment in minority communities have contributed to greater neighborhood disadvantage in Black communities as compared to White^46^, limiting access to educational and economic opportunities, healthy foods, and health care, while increasing exposure to environmental stressors such as air pollution^47–49^. Over time, these cumulative exposures may promote chronic stress and inflammation, accelerating biological aging^50^. Together, our findings suggest that the impact of neighborhood deprivation may be amplified in populations disproportionately exposed to structural disadvantages.

### Strengths and Limitations

Our study has several key strengths. First, we utilized a well-characterized cohort with detailed data on lifetime adversity within a defined geographic region, allowing for stronger comparisons across individuals. Second, we used four well-established DNAm-derived epigenetic clocks, generally observing consistent associations across three of the four even after accounting for multiple comparisons. Third, whereas prior work has largely examined individual or neighborhood factors in isolation^4,34,44,46^, we jointly evaluated neighborhood and individual-level factors. This approach offers a more comprehensive framework to understand the complex interplay between individual and neighborhood-level contributors to the biological aging process.

However, our study has important limitations to consider. Its cross-sectional design prevents casual inference and the ability to assess the impact of long-term exposure to deprivation; thus, mediation analyses should be considered hypothesis-generating. Given sample demographics, we were also unable to assess for race-related effects outside of Black and White participants, and the sample is geographically limited to Connecticut, which may limit generalizability. In addition, ADI, based on census tract-level residence, reflects current residence and does not capture lifetime changes in residence and neighborhood conditions.

Future studies should use longitudinal design to better characterize how individual and neighborhood-level exposures interact over time to shape aging trajectories, considering cumulative neighborhood exposure as prior studies suggest^14,35,51^. Replication in more diverse and nationally representative cohorts is needed to address geographic and demographic limitations in our sample. Finally, expanding measures of the neighborhood environment to include protective factors (e.g., social cohesion, green space) may help identify potential interventions targets beyond education and adversity.

## Conclusion

In this cross-sectional study, greater neighborhood deprivation was associated with accelerated epigenetic aging across multiple second-generation clocks, with effects partially mediated by lifetime adversity and stronger associations observed in Black participants. These results suggest that both neighborhood environments and cumulative psychosocial stress may contribute to biological aging processes and help explain racial disparities in aging trajectories. Future research should prioritize longitudinal studies in more diverse samples to evaluate causal mechanisms and identify neighborhood- and stress-based interventions that will reduce social and racial disparities in epigenetic age acceleration.

## Data Availability

All data produced in the present study are available upon reasonable request to the authors.

https://www.neighborhoodatlas.medicine.wisc.edu/

https://www.census.gov/data/developers/data-sets/acs-5year/2010.html

https://www.icpsr.umich.edu/sites/nanda/view/studies/123001

https://www.researchgate.net/publication/282126949_A_national_spatiotemporal_exposure_surface_for_NO2_monthly_scaling_of_a_satellite-derived_land-use_regression_2000_-_2010

https://www.openicpsr.org/openicpsr/project/141121/version/V3/view

## Conflicts of Interest

The authors declare no conflicts of interest relevant to this study.

## Acknowledgements/Funding

Z.M.H. was supported by the Yale Physician Scientist Development Award and CTSA (NIH UL1 TR001863) and NIDDK grant K23DK136932. Dr. Harvanek is a Pepper Scholar with support from the Claude D. Pepper Older Americans Independence Center at Yale School of Medicine (P30AG021342). This work was supported by NIH grants PL1-DA024859 and UL1-DE19586, as well as NIMH grant K23-MH129684 (B.S.K.).

## Notes

### Competing Interest Statement

The authors have declared no competing interest.

### Funding Statement

This study was funded by NIH grants PL1-DA024859 and UL1-DE19586, as well as NIMH grant K23-MH129684. Z.M.H. was supported by the Yale Physician Scientist Development Award and CTSA (NIH UL1 TR001863) and NIDDK grant K23DK136932. Dr. Harvanek is a Pepper Scholar with support from the Claude D. Pepper Older Americans Independence Center at Yale School of Medicine (P30AG021342).

### Author Declarations

The IRB of Yale University gave ethical approval for this work.

