## Supplementary Materials for "Neighborhood Deprivation Is Associated with Accelerated Epigenetic Aging Via Greater Individual Adversity"

**Supplementary methods:** Cohort recruitment and mediation analysis details

**eTable 1:** Full Model Estimates for Associations between Epigenetic Clocks and the Area Deprivation Index, Individual-level Factors, and Neighborhood-level Factors without Cell Proportions

**eTable 2:** Full model Estimates for Associations between Epigenetic Clocks and Individual-level and Neighborhood-level Factors

**eTable 3:** Full model Estimates for Multivariable Associations between Epigenetic Clocks and Area Deprivation Index, accounting for individual-level and neighborhood-level factors

**eTable 4:** Full model Estimates for Multivariable Associations between Epigenetic Clocks and Neighborhood-Level Factors, excluding the Area Deprivation Index

**eTable 5:** Mediated Effects of Area Deprivation Index on Epigenetic Clocks via Individual-level Factors

**eTable 6:** Full Moderated Mediation Effects of Area Deprivation Index on Epigenetic Clocks via Individual-level Factors

**eTable 6:** Final Moderated Mediation of Area Deprivation Index on Epigenetic Clocks via Individual-level Factors

This supplementary material has been provided by the authors to give readers additional information about their work.

### Supplementary Methods

#### Cohort recruitment details

Participants were recruited through local newspapers, online advertisements, and a local community center between 2008 to 2012. Participants were excluded for psychiatric, medical, or cognitive reasons. Psychiatric exclusions included an active mental health disorder or substance use disorder (excluding nicotine), as determined by the Structured Clinical Interview for Diagnostic and Statistical Manual of Mental Disorders 4th Edition (American Psychiatric Association, 1994), or current use of prescribed psychiatric medications. Medical exclusions include chronic medical conditions (e.g., hypertension, diabetes, hypothyroidism), pregnancy, or prescribed medications for any psychiatric or medical disorders. Cognitive exclusions included inability to read English at a sixth-grade level or higher or a history of head injury with loss of consciousness exceeding 30 minutes. To verify drug abstinence, urine toxicology and breathalyzer screens were administered during each appointment. All participants provided verbal and written informed consent, and the study protocol was reviewed and approved by the Yale Institutional Review Board (IRB). Participants were financially compensated for their involvement in the study.

Prior to enrollment, participants completed two intake sessions to verify eligibility, which included a cumulative stress interview and a morning biochemical evaluation following an overnight fast. Individual-level sociodemographic factors were assessed during the interview, while neighborhood-level variables were derived from geocoded residential addresses and epigenetic aging measures generated from DNA methylation of participant blood samples.

#### Mediation analysis details

Neighborhood disadvantage (ADI rank) was specified as the predictor, lifetime adversity and years of education as parallel mediators, and epigenetic aging indices as outcomes, adjusting for age, gender, and immune cell composition. Indirect effects were estimated using 5,000 bootstrap resamples. Mediation models simultaneously estimated a-paths (ADI→mediators), b-paths (mediators→outcomes), and the direct effect (c′), with total indirect effects computed as the sum across mediators. Moderated mediation models included race (Black=1, White=0) as a moderator of a-, b-, and/or c′-paths via interaction terms. All mediation models were limited to only Black and White participants due to low sample size for other races. Conditional indirect effects were estimated by race, and indices of moderated mediation were calculated as between-group differences. Final models retained only significant moderation pathways for parsimony.

### Supplementary Tables

**eTable 1: Full Model Estimates for Associations between Epigenetic Clocks and the Area Deprivation Index, Individual-level Factors, and Neighborhood-level Factors without Cell Proportions**

In table below, unstandardized effect sizes are presented. P values are nominal; *: p < 0.05; ** p < 0.0125 (Bonferroni correction); *** p < 0.001

| **Predictor** | **GrimAge β (p)** | **PCGrimAge β (p)** | **PhenoAge β (p)** | **PCPhenoAge β (p)** |
| --- | --- | --- | --- | --- |
| Area Deprivation Index | 0.035*** (<.001) | 0.015* (0.042) | NS (0.45) | 0.038** (0.0072) |
| **Individual-level Factors** |  |  |  |  |
| Lifetime Adversity | 0.081*** (<.001) | 0.047*** (<0.001) | NS (0.32) | 0.065** (0.0041) |
| Individual Income | −0.00023* (0.039) | NS (0.11) | NS (0.087) | NS (0.51) |
| Individual Education | −0.30*** (<.001) | −0.19*** (<0.001) | NS (0.096) | −0.29** (0.0020) |
| **Neighborhood-level Factors** |  |  |  |  |
| Food access | 1.2** (.0093) | NS (0.15) | NS (.087) | 1.42** (0.0038) |
| % White Residents | −0.021** (.0054) | NS (0.73) | NS (.90) | NS (0.074) |
| Neighborhood education | −0.040*** (<.001) | −0.027*** (<0.001) | NS (.060) | −0.048*** (<0.001) |
| Neighborhood income | −0.030** (0.0067) | NS (0.24) | NS (.90) | NS (0.16) |
| NO₂ exposure | NS (.058) | NS (.67) | NS (.079) | NS (0.54) |
| Historical redlining | NS (.83) | NS (.79) | NS (.70) | NS (0.69) |

**eTable 2: Full model Estimates for Associations between Epigenetic Clocks and Individual-level and Neighborhood-level Factors after accounting for Cell Proportions**

In table below, unstandardized effect sizes are presented. P values are nominal; *: p < 0.05; ** p < 0.0125 (Bonferroni correction); *** p < 0.001

| **Predictor** | **GrimAge β (p)** | **PCGrimAge β (p)** | **PhenoAge β (p)** | **PCPhenoAge β (p)** |
| --- | --- | --- | --- | --- |
| **Individual-level Factors** |  |  |  |  |
| Lifetime Adversity | 0.080*** (< 0.001) | 0.045*** (< 0.001) | NS (0.47) | 0.060*** (< 0.001) |
| Individual Income | −0.00027* (0.014) | −0.00020** (0.011) | −0.00047** (0.010) | NS (0.72) |
| Individual Education | −0.27*** (<0.001) | −0.15*** (<0.001) | NS (0.32) | −0.19** (0.0063) |
| **Neighborhood-level Factors** |  |  |  |  |
| Historical redlining | NS (0.77) | NS (0.74) | NS (0.81) | NS (0.97) |
| NO₂ exposure | 0.25** (0.0055) | NS (0.212) | NS (0.49) | NS (0.41) |
| Neighborhood income | −0.035*** (<0.001) | −0.017* (0.017) | NS (0.69) | −0.030* (0.013) |
| % White Residents | −0.025*** (<0.001) | NS (0.079) | NS (0.70) | −0.025** (0.0028) |
| Neighborhood education | −0.035*** (< 0.001) | −0.021*** (<0.001) | NS (0.25) | −0.031** (0.0033) |
| Food access | 1.0* (0.014) | NS (0.16) | NS (0.20) | 1.4** (0.0038) |

**eTable 3: Full model Estimates for Multivariable Associations between Epigenetic Clocks and Area Deprivation Index, accounting for individual-level and neighborhood-level factors**

In table below, unstandardized effect sizes are presented. P values are nominal; *: p < 0.05; ** p < 0.0125 (Bonferroni correction); *** p < 0.001

| **Predictor** | **GrimAge β (p)** | **PCGrimAge β (p)** | **PhenoAge β (p)** | **PCPhenoAge β (p)** |
| --- | --- | --- | --- | --- |
| **Individual-level Factors** |  |  |  |  |
| Lifetime Adversity | 0.066*** (< 0.001) | 0.038*** (<0.001) | NS (0.74) | 0.044** (0.012) |
| Individual Income | NS (0.078) | NS (0.07) | −0.00043* (0.021) | NS (0.91) |
| Individual Education | −0.17** (0.0026) | −0.094* (0.027) | NS (0.59) | NS (0.072) |
| Area Deprivation Index | 0.025** (0.0027) | 0.012* (0.037) | NS (0.34) | 0.034** (0.0012) |

**eTable 4: Full model Estimates for Multivariable Associations between Epigenetic Clocks and Neighborhood-level Factors, excluding the Area Deprivation Index**

In table below, unstandardized effect sizes are presented. P values are nominal; *: p < 0.05; ** p < 0.0125 (Bonferroni correction); *** p < 0.001

| **Predictor** | **GrimAge β (p)** | **PCGrimAge β (p)** | **PhenoAge β (p)** | **PCPhenoAge β (p)** |
| --- | --- | --- | --- | --- |
| **Neighborhood-level Factors** |  |  |  |  |
| Food access | NS (0.68) | NS (0.93) | NS (0.14) | NS (0.53) |
| % White Residents | NS (0.95) | NS (0.42) | NS (0.23) | NS (0.40) |
| Neighborhood education | −0.041** (0.0083) | −0.030** (0.0042) | NS (0.074) | −0.038* (0.038) |
| Neighborhood income | NS (0.97) | NS (0.96) | NS (0.28) | NS (0.28) |
| NO₂ exposure | 0.24* (0.036) | NS (0.13) | NS (0.51) | NS (0.55) |

**eTable 5: Mediated Effects of Area Deprivation Index on Epigenetic Clocks via Individual-level Factors**

For mediation analyses: *: p < 0.05; ** p < 0.01; *** p < 0.001

| **Parameter** | **Unstandardized Estimates** | **Standard Error** | **p (>\|Z\|)** | **Standardized Values** |
| --- | --- | --- | --- | --- |
| **GrimAge** |  |  |  |  |
| *A1*: ADI → CSC | 0.102 | 0.035 | .003** | 0.185 |
| *A2*: ADI→ EdYears | -0.025 | 0.008 | .003** | -0.187 |
| B1: CSC → GrimAge | 0.074 | 0.017 | <.001*** | 0.244 |
| B2: EdYears → GrimAge | -0.159 | 0.085 | .062 | -0.125 |
| C: ADI → GrimAge | 0.026 | 0.008 | .002** | 0.153 |
| Indirect: ADI→ CSC→ GrimAge | 0.008 | 0.004 | .03* | 0.045 |
| Indirect: ADI→ EdYears→ GrimAge | 0.004 | 0.002 | .073 | 0.023 |
| Total Indirect Effect | 0.012 | 0.004 | .005** | 0.069 |
| **PCGrimAge** |  |  |  |  |
| *A1*: ADI → CSC | 0.102 | 0.035 | .003** | 0.185 |
| *A2*: ADI→ EdYears | -0.025 | 0.008 | .004** | -0.187 |
| B1: CSC → PCGrimAge | 0.044 | 0.011 | <.001*** | 0.184 |
| B2: EdYears → PCGrimAge | -0.09 | 0.055 | .105 | -0.09 |
| C: ADI → PCGrimAge | 0.012 | 0.006 | .034* | 0.095 |
| Indirect: ADI→ CSC→ PCGrimAge | 0.004 | 0.002 | .032* | 0.034 |
| Indirect: ADI→ EdYears→ PCGrimAge | 0.002 | 0.001 | .119 | 0.017 |
| Total Indirect Effect | 0.007 | 0.002 | .006** | 0.051 |
| **PhenoAge** |  |  |  |  |
| *A1*: ADI → CSC | 0.102 | 0.035 | .003** | 0.185 |
| *A2*: ADI→ EdYears | -0.025 | 0.008 | .004** | -0.187 |
| B1: CSC → PhenoAge | 0.024 | 0.029 | .41 | 0.046 |
| B2: EdYears → PhenoAge | -0.145 | 0.108 | .177 | -0.067 |
| C: ADI → PhenoAge | 0.013 | 0.014 | .336 | 0.047 |
| Indirect: ADI→ CSC→ PhenoAge | 0.002 | 0.003 | .447 | 0.008 |
| Indirect: ADI→ EdYears→ PhenoAge | 0.004 | 0.003 | .232 | 0.013 |
| Total Indirect Effect | 0.006 | 0.004 | .126 | 0.021 |
| **PCPhenoAge** |  |  |  |  |
| *A1*: ADI → CSC | 0.102 | 0.035 | .003** | 0.185 |
| *A2*: ADI→ EdYears | -0.025 | 0.008 | .004** | -0.187 |
| B1: CSC → PCPhenoAge | 0.046 | 0.019 | .017* | 0.105 |
| B2: EdYears → PCPhenoAge | -0.155 | 0.084 | .064 | -0.085 |
| C: ADI → PCPhenoAge | 0.031 | 0.009 | .001** | 0.128 |
| Indirect: ADI→ CSC→ PCPhenoAge | 0.005 | 0.003 | .091 | 0.02 |
| Indirect: ADI→ EdYears→ PCPhenoAge | 0.004 | 0.002 | .068 | 0.016 |
| Total Indirect Effect | 0.009 | 0.003 | .01* | 0.035 |

**eTable 6: Full Moderated Mediation Effects of Area Deprivation Index on Epigenetic Clocks via Individual-level Factors**

For mediation analyses: *: p < 0.05; ** p < 0.01; *** p < 0.001

| **Parameter** | **Unstandardized Est.** | **Standard Err.** | **p (>\|Z\|)** | **Std. all (for figures)** |
| --- | --- | --- | --- | --- |
| **GrimAge** |  |  |  |  |
| Race → GrimAge | 4.013 | 3.333 | .229 | 0.512 |
| Race → CSC | -0.676 | 5.374 | .9 | -0.026 |
| Race → EdYears | -1.658 | 0.854 | .052 | -0.286 |
| Race x ADI → GrimAge | 0.056 | 0.022 | .011* | 0.395 |
| Race x ADI → CSC | 0.118 | 0.107 | .272 | 0.251 |
| Race x ADI → EdYears | 0.007 | 0.019 | .698 | 0.064 |
| Race x CSC → GrimAge | 0.011 | 0.032 | .735 | 0.041 |
| Race x EdYears → GrimAge | -0.353 | 0.207 | .087 | -0.643 |
| **PCGrimAge** |  |  |  |  |
| Race → PCGrimAge | 2.522 | 2.193 | .25 | 0.41 |
| Race → CSC | -0.676 | 5.374 | .9 | -0.026 |
| Race → EdYears | -1.658 | 0.854 | .052 | -0.268 |
| Race x ADI → PCGrimAge | 0.026 | 0.017 | .129 | 0.234 |
| Race x ADI → CSC | 0.118 | 0.107 | .272 | 0.251 |
| Race x ADI → EdYears | 0.007 | 0.019 | .698 | 0.064 |
| Race x CSC → PCGrimAge | -0.002 | 0.023 | .937 | -0.009 |
| Race x EdYears → PCGrimAge | -0.207 | 0.134 | .123 | -0.48 |
| **PhenoAge** |  |  |  |  |
| Race → PhenoAge | 9.556 | 6.856 | .163 | 0.713 |
| Race → CSC | -0.676 | 5.375 | .9 | -0.026 |
| Race → EdYears | -1.658 | 0.854 | .052 | -0.268 |
| Race x ADI → PhenoAge | 0.028 | 0.049 | .57 | 0.114 |
| Race x ADI → CSC | 0.118 | 0.107 | .272 | 0.251 |
| Race x ADI → EdYears | 0.007 | 0.019 | .698 | 0.064 |
| Race x CSC → PhenoAge | -0.063 | 0.068 | .354 | -0.141 |
| Race x EdYears → PhenoAge | -0.613 | 0.439 | .162 | -0.653 |
| Race x CSC → PhenoAge | -0.063 | 0.068 | .354 | -0.141 |
| **PCPhenoAge** |  |  |  |  |
| Race → PCPhenoAge | 5.223 | 3.81 | .17 | 0.463 |
| Race → CSC | -0.676 | 5.373 | .9 | -0.026 |
| Race → EdYears | -1.658 | 0.854 | .052 | -0.268 |
| Race x ADI → PCPhenoAge | 0.058 | 0.025 | .021* | 0.283 |
| Race x ADI → CSC | 0.118 | 0.107 | .272 | 0.251 |
| Race x ADI → EdYears | 0.007 | 0.019 | .698 | 0.064 |
| Race x CSC → PCPhenoAge | -0.041 | 0.039 | .297 | -0.109 |
| Race x EdYears → PCPhenoAge | -0.241 | 0.232 | .299 | -0.305 |

**eTable 7: Final Moderated Mediation of Area Deprivation Index on Epigenetic Clocks via Individual-level Factors**

For mediation analyses: *: p < 0.05; ** p < 0.01; *** p < 0.001

| **Parameter** | **Unstandardized Est.** | **Standard Err.** | **p (>\|Z\|)** | **Std. all (for figures)** |
| --- | --- | --- | --- | --- |
| **GrimAge** |  |  |  |  |
| *A1*: ADI → CSC | 0.102 | 0.035 | .003** | 0.185 |
| *A2*: ADI→ EdYears | -0.025 | 0.008 | .004** | -0.187 |
| B1: CSC → GrimAge | 0.062 | 0.016 | <.001*** | 0.207 |
| B2: EdYears →GrimAge | -0.124 | 0.078 | .111 | -0.099 |
| C: ADI → GrimAge (Race = 0) | -0.002 | 0.009 | .859 | -0.01 |
| C: ADI → GrimAge (Race = 1) | 0.059 | 0.019 | .002** | 0.421 |
| Race x ADI → GrimAge | 0.061 | 0.021 | .004** | 0.431 |
| **PCPhenoAge** |  |  |  |  |
| *A1*: ADI → CSC | 0.102 | 0.035 | .003** | 0.185 |
| *A2*: ADI→ EdYears | -0.025 | 0.008 | .004** | -0.187 |
| B1: CSC → PCPhenoAge | 0.027 | 0.018 | .15 | 0.061 |
| B2: EdYears →PCPhenoAge | -0.085 | 0.066 | .201 | -0.047 |
| C: ADI → PCPhenoAge (Race = 0) | -0.009 | 0.01 | .333 | -0.04 |
| C: ADI → PCPhenoAge (Race = 1) | 0.047 | 0.022 | .032* | 0.239 |
| Race x ADI → PCPhenoAge | 0.057 | 0.024 | .020* | 0.279 |
